# Public but ineffective: Fatality inquiries into childhood deaths in Alberta

**DOI:** 10.1101/2023.09.26.23296196

**Authors:** Mary-Claire Verbeke, Ian Mitchell

## Abstract

**Introduction:** Child Death Review (CDR) processes are public efforts to review a child’s death to understand how and why children die, to improve child health and to prevent future deaths. In 2013, the Canadian Paediatric Society made specific recommendations to establish structured and comprehensive CDR systems in each province and territory. In Alberta, there is no comprehensive CDR process but there are some components. The most public and probably the most expensive component is the public fatality inquiry. A new notification policy adopted after in June 2017 appears to be of limited value for child death prevention.

**Methods:** We examined all Alberta fatality inquiry reports from January 1, 1995 to April 15, 2023 concerning children aged 0-17 years (n=133) to determine whether the fatality inquiry system might be effective in preventing future similar deaths.

**Results:** Recommendations made by judges in a fatality inquiry were not always followed by action, and hence inquiry recommendations have been largely ineffective. Fatality inquiry recommendations were sometimes untimely, and therefore had little chance of being effective. There is an increasing trend from 1995 (case 1) to 2023 (case 133) in the time taken to initiate a child fatality inquiry review.

**Discussion and Conclusion:** Information and recommendations from fatality inquiries into Alberta childhood deaths tend to be delayed and not followed by action. A comprehensive CDR process is required in Alberta. With system changes, public fatality inquiries could be an effective part of the child death prevention process.

## INTRODUCTION

Child death reviews (CDR), at their best, can determine the cause and manner of a child’s death, determine whether there is a need for public protection, and help prevent future similar deaths.^1^ The Canadian Paediatric Society (CPS) has recommended comprehensive, standard, legislated and structured CDRs for all jurisdictions.^2^ A 2016 review showed that such a standard process was lacking in seven jurisdictions (Alberta, Saskatchewan, Nova Scotia, Prince Edward Island, Yukon Territory, Northwest Territory, and Nunavut).^3^

In Alberta, there is no comprehensive CDR, but nevertheless some deaths are reviewed.[2] For example, in most facilities that care for children, the review of all deaths is part of quality improvement and quality assurance programs. There are two legislated reviews of death: that by the Child and Youth Advocate^4^ which is specific to children; and those by the Alberta Office of the Chief Medical Examiner (OCME).^5^ The OCME, by the authority of the *Fatality Inquiries Act*^*6*^ must determine and report accurately all non-natural deaths and some natural deaths. These include all deaths that are sudden and unexplained, with specific sub-categories contained within the Act. The OCME role is to determine the identity of the deceased; date and place of death; medical reason for death; and manner of death. OCME investigate deaths falling under their jurisdiction in individuals of all ages, including children. The Alberta Fatality Review Board is an independent panel that reviews deaths investigated by the OCME in the following circumstances: when the cause and manner of death have not been established; a person dies in a correctional facility, or in the custody of a peace officer, or from use of force by an on-duty peace officer; a patient dies while detained under the Alberta *Mental Health Act*^*7*^ or in a government facility; the Chief Medical Examiner considers a review of the investigation to be necessary; or a review is requested by the next of kin.^8^ In addition, a review is conducted by the Fatality Review Board when “a child dies under the province’s guardianship or in its custody”.^9^ The Board may recommend to the Alberta Minister of Justice and Solicitor General that a public fatality inquiry be held into someone’s death. The Minister makes the order for a public fatality inquiry,^10^ which is held before a Provincial Court Judge. The judge must make a written public report, which may or may not contain recommendations to prevent future similar deaths. There is no requirement that an inquiry transcript be available.

When a judge makes recommendations for change, a government agency or department arguably ought to implement such action. A November 2016 Fatality Inquiry Report recommended that the Government of Alberta “prepare an implementation report on the status of implementation of recommendations from all fatality inquiries.”^11^ In response to this recommendation, the Alberta government established a tracking system to follow and to review inquiry recommendations.^12^ Since apparently June 2, 2017^13^ with the report into the death Marc Fontaine, the Fatality Inquiries website has contained letters from the Fatality Inquiry Coordinator notifying relevant government departments of specific recommendations made by the judge. The site also contains the department’s response, if any.

We aimed to determine whether Alberta public fatality inquiries in the case of childhood deaths published in the period, January 1, 1995 to April 15, 2023, were useful in preventing future similar deaths of children. It is not possible to know in individual cases if prevention was successful. Deaths that did not occur cannot be determined! Therefore, we sought evidence that the reviews were conducted in a timely fashion such that the inquiry might possibly prevent subsequent similar deaths, whatever might be learned from an inquiry was communicated in a recommendation or recommendations, and that recommendations made have been followed.

## METHODS

We reviewed all Alberta public fatality inquiry reports published between January 1, 1995 and April 15, 2023 on the Alberta Justice and Solicitor General Ministry (“the Ministry”) website concerning people younger than age 18. There were 133 such deaths. On a spreadsheet, we listed for each death: the name, sex and age of the deceased (if so stated); the cause, manner, and circumstance of death; the date the inquiry commenced and the date the report was issued, whether the report made recommendations to prevent future such deaths, and whether there were letters communicating the recommendations to relevant departments and agencies and the department or agencies’ response, if any. We then identified whether circumstances amenable to prevention identified in previous reports in the same dataset were present. We also reviewed the responses - there were 101 - by the Ministry or other governmental and non-governmental entities to the Inquiry recommendations. We sought to identify whether items identified as important in the recommendations, appeared in subsequent public fatality inquiry reports.

We grouped our findings into areas that had medical causes of death and manners of death in common and that seemed important for child death prevention. For each of these areas we identified at least one example.

## RESULTS

### 1. Recommendations made too late to prevent subsequent deaths

This analysis revealed that fatality inquiry recommendations were sometimes made after a long period of time had elapsed such that a second, third and fourth death from the same cause occurred before lessons from the first death were expressed in recommendations for prevention. For example, in 2010, a 26-week-old baby, named Phoenix Majestic Omeasoo, died in foster care from “positional asphyxia” while sleeping. The judge’s report from the fatality inquiry into the death of this child was published four years later, in 2014.^14^ The judge recommended that the Alberta Department of Human Services research, identify, and inform foster parents of the best practices for the use of sleeping arrangements for children; such practices include “back to sleep”.^15^ During that four-year interval prior to publication of the Phoenix Omeasoo inquiry report, three other children died in a similar manner. A second child, 16 week old Delonna Victoria Sullivan, died in April of 2011, in her crib with no determined cause. No recommendations were made. The judge stated that “the suggestions from the scientific and medical communities aimed at reducing the risk of this tragic occurrence are already in place in the Children & Youth Services’ ‘Safe Sleep’ policies, procedures and training.”^16^ A third child, Mya Shiningstar-Bird, died at 13 weeks in September of 2011 in foster care; she was found face down in her crib.^17^ No recommendations were made, yet expert witnesses stated that babies should sleep on their backs, and should not share a sleep surface with an adult. A fourth child, named Anthony James Leadly, died in foster care of a similar cause in May of 2011; the 14-week-old boy was found face down, deceased in his crib.^18^ The inquiry into this 2011 death led to the judge’s recommendation that the Alberta Department of Human Services produce a short film demonstrating safe sleep practices.^19^ The judge wrote,

> Because practices involving infant sleeping positions have varied significantly over time, it is recommended that all caregivers, including grandparents for example, be shown the film or be instructed to view it online or through post-natal community care.

The judge noted that such a film would benefit all children, not just children in foster care.

### 2. Inquiries are taking an increasing amount of time to commence and to complete

Our second finding is linked to the first above: there is an increasing trend from 1995 (case 1)^20^ to 2023 (case 133)^21^ in the time taken to initiate a child fatality inquiry review.

(Figure 1) The least amount of time occurred in 1996 (case 7)^22^ -– 138 days – and the longest amount of time (case 117)^23^ was 3755 days (Figure 2). Case 117 is the 2017 inquiry into the death of Aminat Magomadova that occurred in 2007. The amount of time between the child’s death and the start of the inquiry increased in the period under study. Additionally, the amount of time from the commencement of an individual inquiry to the completion of the judicial report also steadily increased.

**Figure 1:**
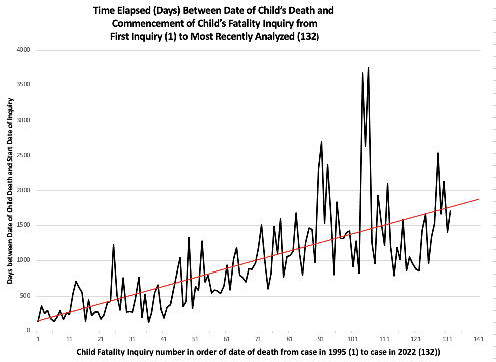
Time Elapsed (Days) Between Commencement to Conclusion of Child’s Fatality Inquiry from First Inquiry (1) to Most Recently Analyzed (133)

**Figure 2:**
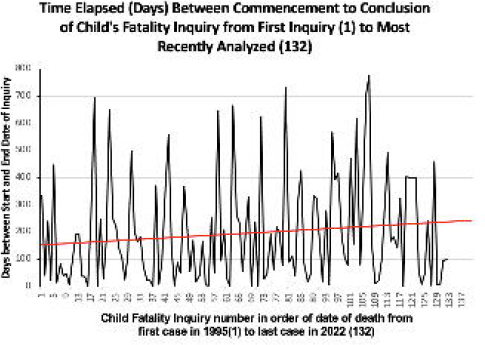
Time Elapsed (Days) Between Date of Child’s Death and Commencement of Child’s Fatality Inquiry from First Inquiry (1) to Most Recently Analyzed (133)

### 3. Multiple inquiries over time are repeating similar recommendations

A third finding, related to increasing time between time of death and report is that numerous cases of similar deaths were followed by inquiries that made similar recommendations. For example, on ten occasions, judges recommended that medical and other files be shared and information exchanged among workers of the Ministry of Children’s Services and with other departments and healthcare facilities. Of the cases we reviewed, the first recommendation was made in 1997 after the death of a female teenager named “O.R.C.” in foster care in 1995.^24^ The girl had a history of depression and suicidal ideation; physicians recommended that she be placed in a secure treatment facility. Instead, a child welfare worker, unfamiliar with the girl’s case and without the benefit of a current medical file, placed the girl in a home in which she died of morphine toxicity. In the 1997 fatality inquiry report regarding her death, the judge recommended that no child be placed in foster care without the proper documentation accompanying the child, and without a formal intake conference involving an informed member of the Department of Family and Social Services and the prospective caregiver.^24^ Had the recommendations made been approved and implemented in a timely manner, then perhaps nine subsequent children would have lived.

### 4. Approximately one third of judicial recommendations are rejected or ignored

The June 2017 change that brought about the notification policy has potential to make a difference. That change occurred when a judge asked inquiry council to determine whether anyone had died in circumstances similar to those before the judge. When the answer was found to be yes, Judge Rosborough made the recommendation that the Government of Alberta prepare an implementation report on the status of implementation of recommendations from all fatality reports.^25^ The Judge’s comments stress the importance of accountability of fatality inquiries to be effective in preventing future death by the same cause,^25^

> Fatality inquiries are costly endeavors […] and while it is for the Government of Alberta to determine whether any or all of these recommendations should be implemented, it is important for community agencies, the media and members of the public to know what action (if any) has been taken pursuant to those recommendations.

Some departments and agencies have responded positively and relatively quickly when notified of a fatality inquiry recommendation. For example, regarding an inquiry into the suicidal death of a child who sat on the railway tracks, the Royal Canadian Mounted Police (RCMP) were notified on January 28, 2022 of an inquiry recommendation concerning the RCMP’s ability to communicate rapidly. The Alberta RCMP responded three months later, on April 21, 2022, by reporting that they had integrated the emergency numbers of the CP and CN Rail dispatch operators into the RCMP communication system such that RCMP communication operators could now connect immediately with the CP or CN Rail Emergency Dispatch Centre.^26^

Yet not all agencies respond relatively rapidly and positively to notification of inquiry recommendations. Figure 3 summarizes the 101 responses of the associated organizations to recommendations made by a fatality inquiry judge. These responses are categorized as “accepted”, “accepted in principle”, and “not accepted”, among other responses. The largest portion of responses (36%) were accepted in principle, in other words, not as the recommendations were written by the judge. Only 33% of the recommendations were accepted as they had been written, and almost one third (31%) of judicial recommendations received either no response, or the recommendations were rejected.

**Figure 3:**
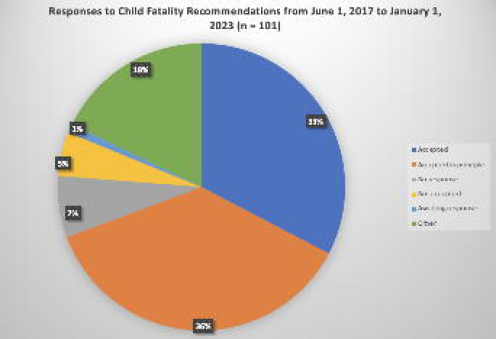
Responses to Child Fatality Recommendations From 2017 (n=101) to April 15, 2023.

These findings reveal that the public inquiry process did not occur in a timely manner such that future similar deaths might have been prevented, and many recommendations made have not been followed by meaningful action, and therefore could not be effective.

## DISCUSSION

We support the Canadian Pediatric Society’s recommendation for CDR systems that are unitary and effective. (27 https://cps.ca/en/documents/position/importance-of-child-and-youth-death-review) In Alberta, one form of death review, the fatality inquiry, is of the longest standing, is the most public and is probably the most expensive of the existing child death reviews. We have noted that on many occasions, that process has been ineffective in preventing future similar deaths and offered specific examples. We believe that if initial recommendations, never mind the many subsequent similar recommendations, had been followed, some of children whose deaths led to fatality inquiries might be alive today.

We believe there are four problems with the fatality inquiry review system. First, there is an inherent delay. The inquiry can occur only after: 1. the police and the OCME have completed their investigations; 2. courts have resolved any related criminal charges, including appeals; and 3. a pre-inquiry conference is completed. Second, only about one third of judicial recommendations are accepted as written and about one third are rejected, raising significant questions as to whether the fatality inquiry process can protect children from future similar death, especially given its high cost. Third, fatality inquiries concerns a small minority of children, albeit a very vulnerable minority, and does not deal with most child deaths. Fourth, there is no requirement to provide a transcript of the hearing which would assist in public understanding of the cause and manner of death and methods to prevent similar deaths.

We would prefer a comprehensive CDR process to be developed in Alberta, with fatality inquiries perhaps being one component but with a different focus and perhaps a reduction in number of inquiries, at least as far as children are concerned.

However, absent a political commitment to a comprehensive CDR process, we assume that this expensive method of child death investigation, the fatality inquiry, will continue. We therefore make the following five recommendations for improvement. First, an electronic file-transfer system might well address the concern that the lack of medical file transfer leads to inappropriate placements of children in care. Second, efforts must be made to allow fatality inquiries to occur in a timely fashion: currently, they take place and publish recommendations years after the child’s death. In that time, more children can die of the same preventable cause. We recognize the priority of maintaining confidentiality of investigation results to avoid tainting criminal processes and see no easy way to reduce delays due to this factor. Nevertheless, other factors that cause delay should still be addressed. Third, inquiries should uniformly include information such as sex, gender, ethnicity, parental background and history of mental illness or substance abuse of the child and of the parent(s), etc. that could be collected into a statistical database. Such a database could help quantify areas requiring the greatest attention (e.g., high suicide rates among adolescents in First Nations communities). Fourth, we recommend that members of the Fatality Review Board have experience, qualifications, and training in fatality investigations. Finally, it must be legally required for all inquiries to make a recommendation to prevent future deaths and for such recommendations to be communicated to the relevant departments and agencies, whose responses (or failure to respond) must be reported publicly.

Some important issues regarding protecting children cannot be addressed even if the fatality inquiry process is improved. Fatality inquiries tend to focus on protecting children in state care, something we support, but they do not provide protections for ALL children. We do not suggest that inquiries should necessarily be public. But we believe that all child deaths should be investigated. Such a change would be easy to effect; the Manitoba *Fatality Inquiries Act* [9] stipulates that all child deaths must be reviewed by using the simple and effective phrase in the statute, “When the deceased is a child.” This short and simple phrase in the governing statute requires that all child deaths be reviewed with fatality inquiries perhaps being one component but with a different focus and perhaps a reduction in number of inquiries, at least regarding children. Therefore, we reiterate our preference for a comprehensive CDR process to be developed in Alberta, and note that this change could be effected simply by the following recommended addition to Section 10 of the Fatality Inquiries Act: “s.10(2)(k) when the deceased is a child”.

## CONCLUSION

Alberta has a public fatality inquiry system, involving people of all ages. In respect of children, we see the inquiry system as part of a disjointed and incomplete approach to childhood deaths. We have made some suggestions regarding how the existing system of fatality inquiries might be improved. Our comprehensive review of Alberta Fatality Inquiry reports might be repeated in other provinces by researchers. We suspect that such research would reveal similar outcomes: an expensive, lengthy system with low effectiveness in preventing child death and does not address all child death. Despite advancing fatality inquiry improvement suggestions, we believe that a fatality inquiry system can never replace a comprehensive CDR process that would be relevant for all children, and might prevent some future deaths.

## Data Availability

All data produced in the present study are available upon reasonable request to the authors.

https://www.alberta.ca/fatality-inquiries

## ACKNOWLEDGEMENTS

We thank Alberta Children’s Hospital Foundation Research Institute for summer studentship funding.

## References

1 Ornstein, A., Bowes, M., Shouldice, M., & Yancha, N. L. (2013). The importance of child and youth death review. Paediatrics & Child Health, 18(8), 426–429.

2 Canadian Paediatric Society. (n.d.). [Internet]. Importance of child and youth death review. Available from: https://cps.ca/en/documents/position/importance-of-child-and-youth-death-review

3 Saskatchewan Prevention Institute. (2016). Child Death Review in Canada: A National Scan [Internet]. Child Death Review in Canada: A National Scan | Canadian Child Welfare Research Portal. [cited January 15, 2023]. Available from: https://cwrp.ca/sites/default/files/publications/en/2-460_child-death-review-in-canada-a-national-scan.pdf

4 Office of the Child and Youth Advocate. (n.d.). What is the OCYA? [Internet]. Available from: https://www.ocya.alberta.ca/ retrieved on 2023-04-12.

5 Office of the Chief Medical Examiner “How the office works, death investigation process, body transportation and report a death.” https://www.alberta.ca/office-chief-medical-examiner.aspx

6 Fatality Inquiries Act, RSA 2000, c F-9, <https://canlii.ca/t/54wrl> retrieved on 2023-04-11.

7 Mental Health Act, RSA 2000, c M-13, <https://canlii.ca/t/55pds> retrieved on 2023-04-11.

8 Government of Alberta, Fatality inquiries, Overview, Fatality Review Board, https://www.alberta.ca/fatality-inquiries.aspx, retrieved on 2023-04-11.

9 Government of Alberta, Fatality inquiries, Overview, Fatality Review Board, https://www.alberta.ca/fatality-inquiries.aspx, retrieved on 2023-04-11.

10 Fatality Inquiries, Act, s. 35(1).

11 The Honourable B.D. Rosborough, Presiding, Provincial Court Judge, Recommendation 7, Alberta Fatality Inquiry Report into the Death of Valerie Wolski, November 10, 2016, Camrose Alberta, https://open.alberta.ca/dataset/cb1046cc-948e-4ab8-a149-3777d83413b7/resource/da313098-712b-4f17-a6ed-d60f7639c943/download/fatality-report-wolski.pdf, retrieved, 2023-04-10.

12 Electronic Mail Correspondence with Abid Mavani, Fatality Inquiry Coordinator, Legal Services Division, Alberta Ministry of Justice, March 10, 2023, regarding a Fatality Inquiry Office change in policy adopted after the November 10, 2016 Fatality Inquiry Report concerning the death of Valerie Wolski.

13 Report to the Minister of Justice and Solicitor General: Public inquiry into the death of Marc Andre Fontaine, June 7, 2017, https://open.alberta.ca/publications/fatality-inquiry-2017-06-07, retrieved 2023-04-10.

14 The Honourable John T. Henderson, Inquiry into the Death of Phoenix Majestic Omeasoo October 3, 2014 at Edmonton, https://open.alberta.ca/dataset/edbdac53-e7fd-4a69-b988-8548648fae06/resource/43438cf2-f9fc-4874-b7d5-c0370fdd7a00/download/2014-fatality-report-phoenix-omeasoo.pdf, retrieved 2023-04-10.

15 Ibid.

16 The Honourable S. M. Bilodeau Inquiry into the Death of Delonna Victoria Sullivan, July 10, 2015 at Edmonton, https://open.alberta.ca/dataset/e67e51ad-2c79-420c-ac96-1cb7eb6be04d/resource/ed549e80-821f-4562-9adc-28a4ff0e1020/download/2015-fatality-report-sullivan.pdf, retrieved 2023-04-10.

17 The Honourable Karim Jivraj, Inquiry into the Death of Mya Shiningstar-Bird, August 14, 2015 at Calgary, https://open.alberta.ca/dataset/db267af0-6044-4cd0-a311-1a56029194e3/resource/abc1ba61-4f79-44cc-8ba0-6dea6494a6ff/download/2015-fatality-report-shiningstar-bird.pdf, retrieved 2023-04-10.

18 The Honourable Mark Tyndale, Inquiry into the Death of Anthony James Leadly, September 15, 2015 at Calgary, https://open.alberta.ca/dataset/2296beb4-6021-494d-ac4e-c11f36d4900c/resource/a094eac8-cbd8-4d3c-abb7-deb5205777ac/download/2015-fatality-report-leadley.pdf, retrieved 2023-04-10.

19 Ibid.

20 The Honourable D.L. Crowe, Inquiry into the Death of Korey Gerlach, January 22, 1996, https://open.alberta.ca/publications/fatality-inquiry-1996-01-22, retrieved 2023-04-10.

21 The Honourable B.R. Hougestol, Inquiry into the death of Haley Marie Penney, February 17, 2023, https://open.alberta.ca/dataset/c65450ce-c350-408b-b3db-4ec25613c0f0/resource/bd9ecd86-46d9-44e7-9915-5121545d1ecc/download/jus-public-fatality-inquiry-2023-02-17.pdf, retrieved 2023-04-10.

22 Inquiry into the death of Skylar Waquan, July 18, 1997, https://open.alberta.ca/dataset/a0c8e10c-33ab-492c-a48c-14295934b808/resource/479dca9c-1e28-41b4-aeb7-daeff5ff8c29/download/00931-report-into-death-of-skylar-waquan.pdf, retrieved 2023-04-10.

23 The Honourable G. Sean Dunnigan, Inquiry into the death of Aminat Magomadova, undated, Filed May 2019, https://open.alberta.ca/dataset/a0c8e10c-33ab-492c-a48c-14295934b808/resource/479dca9c-1e28-41b4-aeb7-daeff5ff8c29/download/00931-report-into-death-of-skylar-waquan.pdf, retrieved 2023-04-10.

24 The Honourable S.A. Hamilton, Inquiry into the death of O.R.C, July 30, 1997, https://open.alberta.ca/dataset/175208a8-93c6-4c39-aa89-38cdbb0f2645/resource/5bed904a-8b7c-41d7-af33-29c5607328c8/download/00932-report-into-death-of-orc.pdf, retrieved 2023-04-12.

25 The Honourable B.D. Rosborough, Presiding, Provincial Court Judge, Recommendation 7, Alberta Fatality Inquiry Report into the Death of Valerie Wolski, November 10, 2016, Camrose Alberta, https://open.alberta.ca/dataset/cb1046cc-948e-4ab8-a149-3777d83413b7/resource/da313098-712b-4f17-a6ed-d60f7639c943/download/fatality-report-wolski.pdf, retrieved, 2023-04-10.

26 Brian Brennan, Deputy Commision, Contract and Indigenous Policing, RCMP, to the Alberta Office of the Deputy Minister of Justice and Deputy Solicitor General, April 21, 2022, https://open.alberta.ca/dataset/a43fb2ab-dd46-453f-892e-afc93aedd625/resource/029c0970-3d29-46f0-882f-ef5a996e5f52/download/jsg-fatality-inquiry-2022-01-17-response2-from-rcmp.pdf

